# Medicare Payment for Calcium Modification Technologies Among Patients Undergoing Percutaneous Coronary Intervention, 2021-2022

**DOI:** 10.1101/2025.01.12.25320423

**Authors:** Jamie Schlacter, Huihui Yu, Sarah Tsuruo, Jeph Herrin, Joseph S. Ross, Leora I. Horwitz, Sanket S. Dhruva

## Abstract

**Background:** The Centers for Medicare and Medicaid Services (CMS) New Technology Add-on Payment (NTAP) program supports adoption of new, costly medical technologies demonstrating substantial clinical improvement. In 2021, CMS waived the “substantial clinical improvement” criterion for devices designated under the FDA Breakthrough Devices Program (BDP). This study characterized risk-standardized payments associated with hospitalizations in which Medicare beneficiaries received calcium modification during PCI for acute myocardial infarction (AMI) following the adoption of the Shockwave C^2^ Coronary Intravascular Lithotripsy (IVL) Catheter (Shockwave Medical) with BDP designation.

**Methods:** We analyzed Medicare beneficiaries hospitalized for AMI who underwent PCI between January 2021 and December 2022, stratifying them into four groups: no calcium modification, rotational atherectomy (RA), orbital atherectomy (OA), and coronary IVL. Risk-standardized Medicare payments at 30 days, including index facility, physician, and post-acute care costs, were assessed using non-parametric median and chi-square tests.

**Results:** Among 87,238 patients, 76,462 (87.6%) received no calcium modification, 8,316 (9.5%) underwent RA, 793 (0.9%) underwent OA, and 1,668 (1.9%) underwent IVL. IVL use increased from 1.6% in October 2021 to 4.4% in December 2022. Median total risk-standardized Medicare payments were significantly higher for patients receiving calcium modification technologies ($27,579 for IVL, $27,353 for OA, $23,240 for RA) compared to those without ($19,115; p<0.001). Payment differences were largest for index facility payments.

**Conclusion:** Coronary IVL during PCI for Medicare patients hospitalized for AMI was associated with significantly increased Medicare payments. Further studies must determine whether IVL, and calcium modification technologies in general, improve outcomes for patients hospitalized for AMI undergoing PCI and thus warrant higher payments via NTAP.

## INTRODUCTION

Since 2001, the Centers for Medicare and Medicaid Services (CMS) New Technology Add-on Payment (NTAP) program has provided supplemental hospital reimbursement for new and costly technologies that offer substantial clinical improvement for Medicare beneficiaries over existing treatment. Orbital atherectomy (OA) is one such technology, approved for NTAPs in 2013 for calcium modification during percutaneous coronary intervention (PCI), over 20 years after the Food and Drug Administration (FDA) approved a predecessor for calcium modification, rotational atherectomy (RA). In 2021, CMS waived the criterion requiring “substantial clinical improvement” for devices authorized after receiving FDA Breakthrough Devices Program (BDP) designation. The BDP provides expedited FDA review and accepts a greater extent of uncertainty of benefits and risks for medical devices that are intended to treat irreversibly debilitating or life-threatening conditions, contingent on adequate postmarket evaluation. Shortly thereafter, FDA approved another calcium modification technology, the Shockwave C^2^ Coronary Intravascular Lithotripsy (IVL) Catheter (Shockwave Medical)^1^ with BDP designation. As costs of these technologies have not been described, we characterized risk-standardized payments associated with hospitalizations in which Medicare beneficiaries received calcium modification during PCI for acute myocardial infarction (AMI).

## METHODS

We identified all patients who underwent PCI during hospitalization for AMI from 1/1/2021 to 12/31/2022. We excluded patients lacking cost data and receiving procedures with ≥2 calcium modification technologies during the index hospitalization. We stratified patients into four groups: no calcium modification technology, RA, OA, and coronary IVL. We estimated risk-standardized Medicare payment at 30 days, an episode-of-care measure that comprises index facility payment, index physician payment, and post-acute care payment and accounts for patient characteristics.^2^ Non-parametric median and chi-square tests were conducted; a p-value <0.05 was significant. The study was approved by the Yale School of Medicine institutional review board.

## RESULTS

Among 218,475 patients hospitalized with AMI in 2021-22, 100,273 underwent PCI. After excluding 507 patients who received multiple calcium modification technologies and 12,528 patients without available cost data, 87,238 (39.9%) patients remained in the study cohort. This cohort had a mean (±standard deviation) age of 76.2 (±7.1) years and were predominantly male (60.4%) and white (88.5%) (**Table**). Overall, 76,462 (87.6%) received no calcium modification technology, 8,316 (9.5%) underwent RA, 793 (0.9%) underwent OA, and 1668 (1.9%) underwent IVL. IVL monthly usage increased from 1.6% in 10/2021 (first introduction of the billing code) to 4.4% in 12/2022. Among patients receiving calcium modification, those receiving coronary IVL (77.4±7.1) years or undergoing OA (77.8±7.1) years were older than patients undergoing RA (75.8±7.0) years. Patients undergoing coronary IVL were slightly less likely to be male (63.5%) than those undergoing RA (64.6%) and OA (63.9%). Those who received coronary IVL were more likely to have a history of prior PCI (31.2%) than those undergoing RA (22.1%) or OA (25.3%).

**Table.**
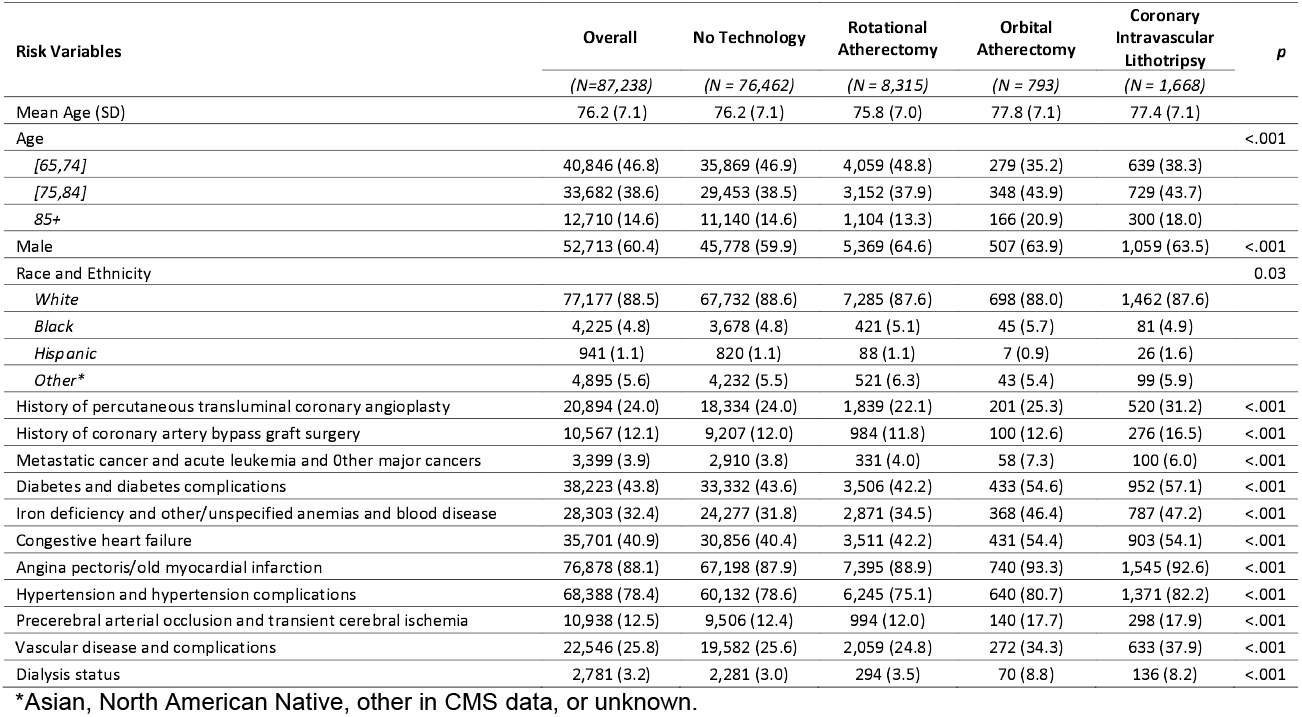
Characteristics of Medicare beneficiaries hospitalized for acute myocardial infarction from 2021 through 2022 categorized by calcium modification technologies used during percutaneous coronary intervention.

The median total risk-standardized Medicare payment was lowest for patients without calcium modification ($19,115, IQR, $15,279-$27,090) compared to those receiving a calcium modification technology (RA: $23,240, IQR, $15,722-$35,430; OA: $27,353, IQR, $19,979-$50,735; coronary IVL: $27,579, IQR, $19,777-$40,263; **Figure**, p<0.001). Payment differences were largest for index facility payments and marginal for index physician and post-acute care payments.

**FIGURE.**
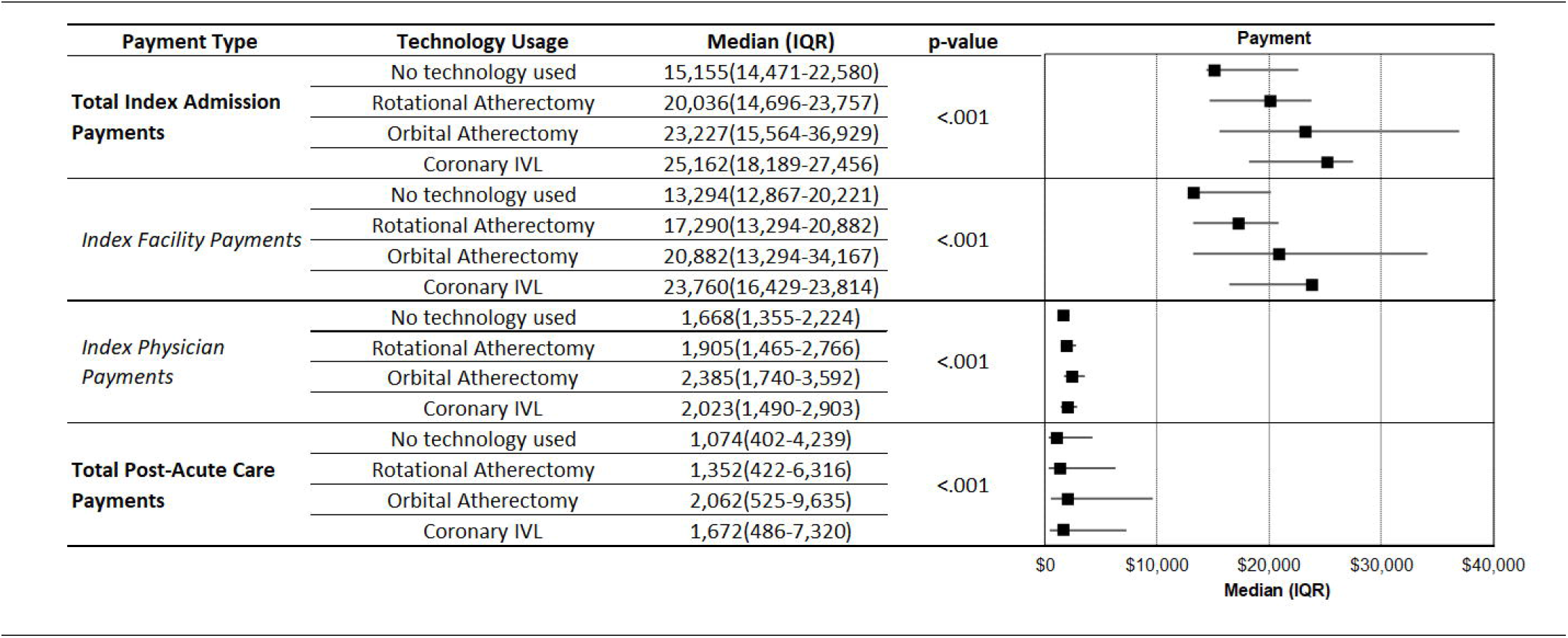
Risk-standardized Medicare payment associated with hospitalization for acute myocardial infarction from 2021 through 2022 categorized by calcium modification technologies used during percutaneous coronary intervention.

## DISCUSSION

Overall, 2% of Medicare beneficiaries hospitalized for AMI received IVL during PCI from 2021-2022. However, monthly IVL use more than doubled over this period, and associated inpatient Medicare payments were nearly twice those of patients not receiving calcium modification technologies, while post-acute care costs remained unchanged. Although heavy coronary calcification is associated with worse stent-related outcomes among patients undergoing PCI,^3^ there is limited evidence that PCI with IVL leads to improved clinical outcomes.^1^ Because IVL’s substantially higher payments resulted from its automatic qualification for the NTAP program due to BDP designation,^1,4^ these findings raise concerns that revenue considerations could be a driver of technology use. In fact, in October 2023, CMS created new Medicare Severity Diagnosis Related Group (MS-DRG)s for patients undergoing PCI with IVL; payment rates are approximately $8,000 higher.^5^

Our study may not generalize to populations treated with PCI for other indications. Additionally, hospitalization costs could be related to peri-procedural factors other than calcium modification strategy; however, the payment measures are risk-adjusted. Finally, RA can be used for reasons other than preparing calcified coronary lesions.

Coronary IVL during PCI for Medicare patients hospitalized for AMI was associated with significantly increased Medicare payments. Further studies must determine whether IVL, and calcium modification technologies in general, improve outcomes for patients hospitalized for AMI undergoing PCI and thus warrant higher payments via NTAP and/or MS-DRG.

## Data Availability

All data used in this study can be requested from the Centers for Medicare and Medicaid Services.

